# ASSOCIATION BETWEEN SLEEP REGULARITY AND SYMPTOMS OF ANXIETY AND DEPRESSION IN MIDDLE-AGED ADULTS

**DOI:** 10.64898/2026.01.15.26344207

**Authors:** Samir Chalise, Laura Nauha, Raija Korpelainen, Maisa Niemelä, Vahid Farrahi

## Abstract

**Background:** Sleep regularity plays a significant role in mental health, but its association with anxiety or depression symptoms in midlife and whether movement behaviors moderate these associations is unclear. We examined how variability in sleep timing and duration is associated with these symptoms and tested the moderating effects of physical activity and sedentary time.

**Methods:** We analyzed data from 3,556 46-year-old participants in the Northern Finland Birth Cohort 1966. Sleep regularity was derived from 7 days of accelerometer-measured sleep timing (midpoint of sleep period) and sleep duration, expressed as standard deviations and categorized as regular, moderately irregular, or highly irregular. Anxiety and depression symptoms were assessed using the Hopkins Symptom Checklist-25. Associations were examined using multivariable linear regression adjusted for gender, marital status, education, smoking, alcohol use, medication, chronotype, work schedule, and physical activity or sedentary time derived from the same accelerometer data, with moderation tested using interaction terms (B coefficients with 95% confidence intervals).

**Results:** Greater sleep timing irregularity was associated with higher anxiety symptoms (B = 0.025, 95% CI [0.007, 0.043], p = 0.001), and with depressive symptoms only when models included sedentary time (B = 0.035, 95% CI [0.017, 0.053], p < 0.001). Sleep duration irregularity showed no significant associations. Physical activity and sedentary time did not moderate these relationships.

**Conclusions:** Sleep timing regularity was more consistently associated with mental health outcomes, especially anxiety, than sleep duration variability. Maintaining regular sleep timing may support anxiety management in midlife, highlighting its potential relevance for preventive intervention.

## Background

Sleep is a fundamental pillar of health, profoundly influencing physical and mental well-being across a person’s lifespan (Ramar et al., 2021). Traditionally, sleep quantity has received significant attention because of its links to increased risks of diabetes, obesity, heart disease, and depression, as well as cardiovascular and mortality risk (Cappuccio et al., 2010; Gallichio & Kalesan, 2009; Shen et al., 2016). However, sleep is recognized as a multidimensional construct that includes not only duration but also the timing and stability of sleep–wake patterns in relation to the circadian system. More recently, emerging research has emphasized sleep regularity as an independent and important predictor of health (Wang et al., 2025; Windred et al., 2023). Sleep regularity refers to the consistency of sleep timing and duration across days, including consistent bedtimes, sleep onset, wake-up times, and sleep period.

Growing evidence supports the association between irregular sleep patterns and unfavorable cardiovascular, cardiometabolic, and cognitive outcomes (Huang et al., 2020; Lunsford-Avery et al., 2018; Nauha et al., 2024; Qin & Chee, 2024; Zuraikat et al., 2020), as well as increased symptoms of anxiety and depression (Howarth & Miller, 2024; Lai et al., 2020; Luo & Lin, 2024; Matcham et al., 2024; Mellman, 2006; Walsh et al., 2025).

Irregularity in sleep timing has been identified as a key disruptor of circadian rhythms and emotional regulation (Phillips et al., 2017; Sletten et al., 2023), underscoring its relevance in both physical and psychological health. Sleep timing regularity indicates the degree of variation in bedtime and wake-up times, which may relate to shifts in biological rhythms and possible effects on mental health (Coelho et al., 2024; Hartstein et al., 2025; Lyall et al., 2018; Wulff et al., 2010; Yin et al., 2023).

Middle-aged adults, typically defined as individuals between 40 and 65 years of age, represent a demographic particularly vulnerable to disrupted sleep patterns due to a confluence of life-stage-specific stressors. This period often includes increased occupational responsibilities, caregiving duties, and age-related physiological changes, all of which can contribute to irregular sleep schedules, such as inconsistent bedtimes and wake-up times (Ohayon et al., 2017). These disruptions can adversely affect mood and emotional regulation, increasing the risk of mental health disorders such as anxiety and depression. Despite the critical importance of this issue, much of the prior research has disproportionately focused on younger or older populations, leaving middle-aged adults relatively understudied in this context (Dregan & Armstrong, 2009; Lunsford-Avery et al., 2018).

Emerging evidence also suggests that lifestyle factors, like physical activity and sedentary time, may moderate the relationship between sleep and mental health (Edwards & Loprinzi, 2017; Kredlow et al., 2015; Zhang et al., 2022). Regular physical activity has been shown to improve mood, reduce symptoms of depression and anxiety, and lower psychological distress (Amiri et al., 2024; Pearce et al., 2022; Singh et al., 2023). Conversely, prolonged sedentary behavior may exacerbate depression and anxiety (Duncan et al., 2023; Edwards & Loprinzi, 2017; Kredlow et al., 2015; Li et al., 2024; Vallance et al., 2011).

While research increasingly associates sleep health with mental health outcomes, current studies lack standardized methods to assess sleep regularity, often relying on subjective measures rather than unobtrusively measured sleep data. Similarly, only a few studies distinguish between variability in sleep timing and sleep duration, and even fewer examine both together (Chaput et al., 2020; Maki et al., 2025). Also, the potential moderating effects of physical activity and sedentary time on these associations have rarely been studied. To address these gaps, this population-based study examined the association between sleep regularity, derived from accelerometer-measured sleep periods, and symptoms of anxiety and depression, and investigated whether physical activity and sedentary time moderated these associations in a large sample of middle-aged adults from the Northern Finland Birth Cohort 1966 (NFBC1966).

## Methods

### Study Population

This study utilized a cross-sectional design and data from the NFBC1966, a prospective, population-based birth cohort originally comprising 12,058 individuals whose stated year of birth was 1966 and who were born in Finland’s northernmost provinces, Oulu and Lapland (Nordström et al., 2021; University of Oulu, 1966). The cohort was investigated at ages 1, 14, 31, and most recently at 46 years (2012–2014). For the latest survey, 10,331 individuals were invited to participate; 7,146 responded to a postal survey, and 5,832 attended clinical examinations. Accelerometer-based 24-hour activity monitoring of 5,621 participants was conducted for 2 weeks. This study included a sample of 3,556 participants from whom 7 consecutive days of valid sleep timing and physical activity data were collected, along with self-reported information on anxiety and depression symptoms. It was approved by the Ethical Committee of the Northern Ostrobothnia Hospital District in Oulu, Finland (94/2011). A flowchart detailing the selection of this study’s population is shown in Figure 1.

**Figure 1.**
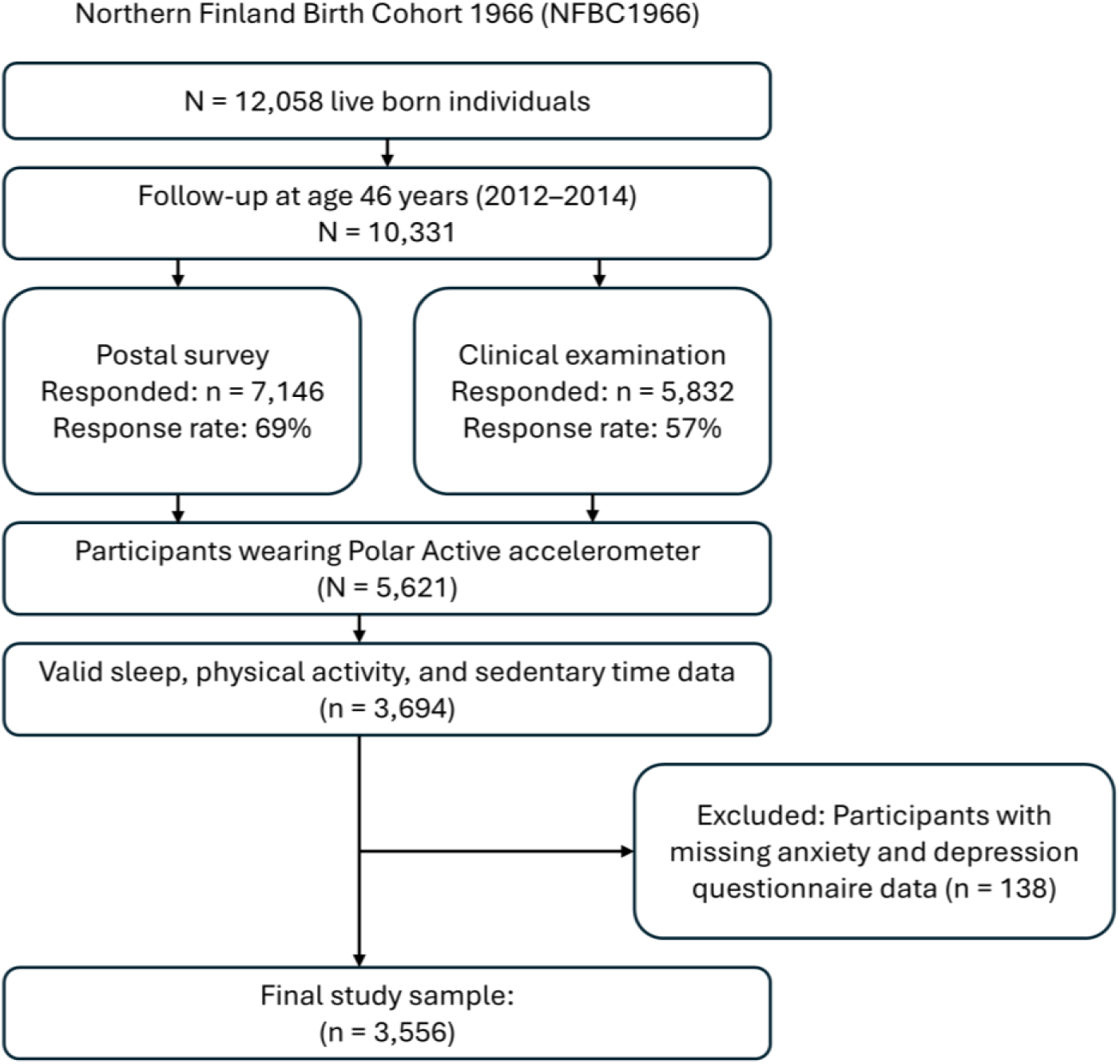
Flow chart of study population

### Measurements

#### Anxiety and Depression Symptoms

The outcome variables in this study were anxiety and depression symptoms, assessed using the Hopkins Symptom Checklist-25 (HSCL-25). HSCL-25 is a validated self-report questionnaire widely used across cultures to measure psychological distress (Nabbe et al., 2019; Sivertsen et al., 2024; Vindbjerg et al., 2021). The HSCL-25 consists of 25 items, of which the first 10 focus on anxiety symptoms (such as headache, difficulty falling asleep, feelings of strain or stress, exhaustion, episodes of panic or anxiety, restlessness, and nervousness), and the remaining 15 focus on depression symptoms (such as low energy, self-blame, crying, sexual dysfunction, feeling despair about one’s destiny, feeling upset, experiencing loneliness, suicidal thoughts, feeling trapped, worrying excessively, feeling no interest in anything, a sense of worthlessness, and reduced appetite; Jacobi et al., 2014; Sivertsen et al., 2024). Each item is rated on a 4-point Likert scale from 1 (*not at all*) to 4 (*extremely*), with higher scores indicating more severe symptoms. The mean symptom scores were calculated as follows: 1) The anxiety symptom score was determined by averaging the 10 anxiety items, and the depression symptom score was calculated by averaging the 15 depression items.

The final symptom scores range from 1.0 to 4.0. The use of a dual-symptoms structure enables both distinct and integrated analyses of anxiety and depression symptom dimensions, allowing for a nuanced understanding of their independent associations with sleep regularity. Additionally, to identify participants with clinically relevant anxiety and depression symptoms, binary categorical variables were created from the continuous HSCL-25 scores.

Based on established clinical thresholds from previous research, participants were categorized as having symptoms of anxiety or depression if their average score on the corresponding subscale was greater than or equal to 1.75, which is a commonly used cut-off indicating potential clinical distress (Derogatis et al., 1974; Housen et al., 2018; Tinghög et al., 2017).

Accordingly, a binary variable for anxiety and depression status was created where 0 = no symptoms (mean score < 1.75) and 1 = symptoms present (mean score ≥ 1.75).

### Accelerometer-Determined Sleep Period, Physical Activity, and Sedentary Time

Sleep period, physical activity, and sedentary time were measured using Polar Active wrist-worn accelerometers (Polar Electro Oy, Kempele, Finland; Kinnunen et al., 2019).

Participants were instructed to wear the accelerometer device on their non-dominant wrist for at least 14 days. The device recorded metabolic equivalents (METs) in 30-second intervals, calculated using background information such as height, weight, age, and sex. Previous validation studies have shown that Polar Active provides an accurate estimate of daily energy expenditure compared to the doubly labeled water method (R² = 0.78; Kinnunen et al., 2019). Participants did not receive any feedback from the device regarding their activity behaviors. A valid day was defined as one with less than 2.5 hours of non-wear time, where non-wear time was defined as continuous periods with intensity levels below 1 MET. Only participants with at least 7 consecutive valid measurement days were included in the analysis.

Sleep periods were derived from Polar Active MET data using a previously validated algorithm that was shown to detect bedtime and wake-up time based on prolonged low-activity periods, identifying bedtime as the start of the low-activity period and wake-up time as its end (Nauha et al., 2023, 2024). The method has been shown to estimate these times with about 20-minute accuracy compared to self-reported sleep diaries and about 5-minute accuracy compared to smart ring data, which utilizes photoplethysmography and acceleration data to detect sleep and shows high validity against polysomnography (de Zambotti et al., 2019).

Both sedentary time and total physical activity were calculated for the hours outside the defined sleep period. Total physical activity included all activity with an intensity of ≥ 2.0 METs, while sedentary time (min/day) was calculated as all time spent with an intensity of 1.0 to 1.99 METs. Total physical activity (MET min/day) was computed by multiplying each MET value (≥ 2.0 METs) by its corresponding duration and then averaging across valid days (Niemelä et al., 2019).

### Sleep Regularity

Sleep regularity was operationalized from sleep period measurements using two variables: sleep timing regularity and sleep duration regularity. Sleep timing regularity was calculated as the standard deviation (SD) of the midpoint of the sleep period across 7 consecutive valid days. This midpoint lies halfway between bedtime and wake-up time. Higher SD values indicate greater fluctuations in sleep timing, reflecting instability in circadian rhythms. Sleep duration regularity was calculated as the SD of the sleep period duration across the same 7 days. Higher SD values indicate greater variability in nightly sleep length.

### Covariates

A range of covariates with independent associations to anxiety or depressive symptoms were controlled for, based on previous literature linking demographic, lifestyle, and sleep-related variables with these outcomes (Alvaro et al., 2013; Edwards & Loprinzi, 2017; Kredlow et al., 2015; Lyall et al., 2018; Wulff et al., 2010; Zhang et al., 2022). Demographic covariates included gender (male vs. female), marital status (living alone vs. married or cohabiting), and education level (basic or secondary vs. higher education).

Lifestyle-related covariates were smoking status (non-smoker/former smoker vs. current smoker), alcohol consumption (calculated to g/day based on responses), sedentary time, and total physical activity. Heavy alcohol drinkers were defined according to the Finnish Institute for Health and Welfare guidelines, which classify heavy drinking as ≥ 40 g/day for men and ≥ 20 g/day for women (Duodecim, 2018). Work-related factors were based on self-reported work schedules and categorized as day work, shift work, and not working/missing data. The use of depression and anxiety medications (yes vs. no) was based on the self-reported use of psychotropic medicines, including antidepressants, anxiolytics, or sleep aids. Chronotype was assessed using the shortened morningness–eveningness questionnaire (MEQ; Hätönen et al., 2008). The MEQ includes six items on preferred times for waking, sleeping, and performing various activities, yielding a score of 5 to 27, classified as evening-type (5–12), day-type (13–18), and morning-type (19–27).

### Statistical Analyses

Sleep regularity was divided into tertiles according to the sleep regularity variable: regular, moderately regular, and highly irregular. This categorization was performed separately for both sleep timing regularity and sleep duration regularity. Descriptive statistics were used to summarize participant characteristics and examine differences across sleep regularity tertiles. Frequencies and percentages were calculated for categorical variables, and chi-square tests were used to assess group differences. For continuous variables, a one-way ANOVA was applied, and means and standard deviations were reported. Anxiety and depression symptom scores (HSCL-25) showed right-skewed distributions; therefore, Kruskal–Wallis tests with Bonferroni-adjusted post hoc comparisons were used for these outcomes.

Multivariable linear regression models were used to analyze the associations between sleep regularity and log-transformed anxiety and depression scores. We evaluated the associations with four incremental models, including an unadjusted model and three adjusted models. The regular sleep group served as the reference category. The unadjusted model included sleep regularity as a categorical variable and outcome variables. Model 1 was adjusted for sociodemographic and lifestyle covariates. Model 2A included all adjustments from Model 1 and additionally physical activity, whereas Model 2B included all adjustments from Model 1 and additionally sedentary time. Regression assumptions were evaluated using residual plots, Q–Q plots, and variance inflation factors (VIF < 1.5). The anxiety and depression symptom variables (HSCL-25 subscale mean scores) were log-transformed to improve normality and homoscedasticity. Missing data were minimal (< 5% for covariates) and were managed via listwise deletion. Interaction terms were used to evaluate whether physical activity or sedentary time moderated associations between sleep regularity and mental health outcomes, and continuous moderators were mean-centered prior to interaction term creation. For each moderator, two interaction terms were created (moderately regular × moderator and highly irregular × moderator, with regular as the reference category).

Continuous moderator variables were mean-centered before creating interaction terms to reduce multicollinearity and improve interpretability. Both main effects and interaction terms were retained in the models to ensure proper estimation of conditional effects. All analyses were conducted using IBM SPSS Statistics version 25.0, with statistical significance set at p < 0.05.

## Results

The final study sample comprised 3,694 middle-aged participants (61% female). Based on the HSCL-25 scores, 9.6% of participants reported clinically relevant anxiety symptoms and 11.8% reported depressive symptoms (HSCL-25 ≥ 1.75). When examining the characteristics of study participants across sleep midpoint regularity groups, those in the regular group had the earliest average sleep midpoint (3:08), bedtimes (22:57), and wake-up times (7:03), as well as the longest sleep period duration (8 h 6 min). In contrast, individuals in the highly irregular group exhibited later sleep timing, shorter sleep period duration (7 h 43 min), lower physical activity, greater sedentary time, and higher anxiety and depression scores (all p < 0.001). Sociodemographic and behavioral differences followed a clear gradient: participants with a highly irregular sleep midpoint were more likely to be single, less educated, current smokers, evening chronotypes, shift workers or unemployed, at higher alcohol risk, and medication users (Table 1).

**Table 1.** Descriptive Characteristics of Study Participants (n = 3,556) Stratified by Sleep Timing Regularity and Sleep Duration Regularity.

| Variable | Midpoint regularity |  |  | p-value | Sleep duration regularity |  |  | p-value |
| --- | --- | --- | --- | --- | --- | --- | --- | --- |
|  | Regular (n=1,185) | Moderately regular (n=1,186) | Highly Irregular (n=1,185) |  | Regular (n=1,185) | Moderately regular (n=1,186) | Highly Irregular (n=1,185) |  |
| Bedtime [hh:mm] | 22:57 (0:55) | 23:11 (1:03) | 23:53 (1:26) | <0.001 | 23:10 (0:58) | 23:17 (1:05) | 23:35 (1:29) | 0.935 |
| Wake-up time [hh:mm] | 7:03 (0:59) | 7:12 (1:03) | 7:35 (1:19) | <0.001 | 7:07 (0:59) | 7:13 (1:05) | 7:31 (1:20) | <0.001 |
| Sleep period [hh:mm] | 8:06 (0:52) | 8:02 (0:56) | 7:43 (1:03) | <0.001 | 8:00 (0:58) | 8:01 (1:05) | 7:56 (1:20) | <0.001 |
| Midpoint [hh:mm] | 3:00 (0:35) | 3:12 (0:32) | 3:43 (1:24) | <0.001 | 3:08 (0:53) | 3:15 (0:58) | 3:32 (1:18) | <0.001 |
| Anxiety score | 1.26 (0.40) | 1.27 (0.40) | 1.33 (0.40) | <0.001 | 1.27 (0.40) | 1.28 (0.40) | 1.32 (0.40) | <0.001 |
| Depression score | 1.34 (0.33) | 1.36 (0.40) | 1.40 (0.47) | <0.001 | 1.35 (0.33) | 1.36 (0.40) | 1.39 (0.40) | <0.001 |
| Physical activity [MET min/day] | 1133 (355) | 1074 (316) | 1055 (328) | <0.001 | 1138 (342) | 1090 (322) | 1034 (332) | <0.001 |
| Sedentary time [min/day] | 564 (93) | 579 (85) | 588 (89) | <0.001 | 574 (89) | 578 (88) | 579 (91) | 0.444 |
| Chronotype, n (%) |  |  |  | <0.001 |  |  |  | <0.001 |
| Morning type | 555 (47.5) | 481 (41.3) | 426 (36.6) |  | 528 (36.1) | 494 (33.8) | 440 (30.1) |  |
| Day type | 573 (49.0) | 624 (53.5) | 642 (55.2) |  | 589 (36.1) | 631 (34.3) | 619 (33.7) |  |
| Evening type | 41 (3.5) | 61 (5.2) | 202 (8.2) |  | 47 (4.0) | 47 (4.0) | 104 (8.9) |  |
| Female, n (%) | 755 (63.7) | 752 (63.4) | 668 (56.4) | <0.001 | 748 (63.2) | 736 (62.1) | 691 (61.2) | 0.37 |
| Current smoker, n (%) | 140 (12.0) | 175 (14.9) | 304 (25.9) | <0.001 | 163 (13.9) | 190 (16.2) | 226 (22.8) | <0.001 |
| Married/cohabiting, n (%) | 968 (82.0) | 954 (80.8) | 856 (72.4) | <0.001 | 965 (81.9) | 956 (80.7) | 857 (72.5) | <0.001 |
| High education, n (%) | 362 (31.6) | 367 (31.9) | 266 (23.6) | <0.001 | 379 (38.1) | 330 (33.2) | 286 (28.7) | 0.002 |
| Heavy alcohol drinkers, n (%) | 59 (5.0) | 89 (7.5) | 126 (10.6) | 0.001 | 74 (6.2) | 82 (6.9) | 118 (10.0) | <0.001 |
| Psychotropic medication use, n (%) | 139 (11.7) | 105 (8.9) | 161 (13.6) | <0.001 | 114 (9.6) | 119 (10.0) | 172 (14.5) | <0.001 |
| Shift work, n (%) | 137 (11.6) | 175 (14.8) | 291 (16.9) | <0.001 | 120 (10.1) | 180 (15.2) | 215 (25.5) | <0.001 |

Values are means (SD) unless otherwise specified. Numbers do not match due to the missing values. Anxiety and depression scores are based on Hopkins Symptom Checklist-25 (HSCL-25) questionnaire. MET: metabolic equivalent. Education is categorized as basic or secondary/higher; table shows proportion with higher education. Heavy alcohol drinkers: men ≥ 40 g/day; women ≥ 20 g/day (Duodecim, 2018) Use of depression and anxiety medication (yes/no) was defined as self-reported use of psychotropic drugs, including antidepressants, anxiolytics, or sleep aids.

When examining the characteristics of study participants across sleep duration regularity groups, the average sleep duration was similar across groups (approximately 7 h 56 min). Although the average duration of the sleep period remained consistent across groups, the highly irregular group exhibited later wake-up times and lower physical activity (both p < 0.001). Those with highly irregular sleep duration were less likely to be married, have higher education, work a day shift, and more likely to smoke, use medications, engage in at-risk alcohol use, and identify as evening chronotypes and shift workers (all p < 0.001, except education at p = 0.002).

Table 2 presents the multivariable regression results for the associations between sleep regularity and HSCL-25 symptoms of depression. In the unadjusted model, participants with highly irregular sleep timing had significantly higher depression scores than those in the regular group (B = 0.038, 95% CI [0.020, 0.056], p < 0.001). Regarding sleep duration regularity, highly irregular duration was also associated with higher depressive symptoms (B = 0.022, 95% CI [0.004, 0.040], p = 0.017). In Model 2B (adjusted for sedentary time), timing irregularity remained associated with higher depression scores (B = 0.035, 95% CI [0.017, 0.053], p < 0.001; Table 2).

**Table 2.** The Association Between Sleep Timing Regularity/Sleep Duration Regularity and Depressive Symptom Scores Among 46-year-old Northern Finland Birth Cohort 1966 Participants (n = 3,556) Using Multivariable Regression Analyses

| Model | Sleep timing regularity |  |  |  |  | Sleep duration regularity |  |  |  |
| --- | --- | --- | --- | --- | --- | --- | --- | --- | --- |
|  | Category | B (95% CI) | Adjusted R<sup>2</sup> | p |  | Category | B (95% CI) | Adjusted R<sup>2</sup> | p |
| **Unadjusted** | Regular (ref.) | – | 0.011 | – |  | Regular (ref.) | – | 0.001 | – |
|  | Moderately regular | 0.012 (–0.008, 0.032) |  | 0.380 |  | Moderately regular | 0.002 (–0.017, 0.020) |  | 0.867 |
|  | Highly irregular | **0.038 (0.020, 0.056)** |  | **<0.001** |  | Highly irregular | **0.022 (0.004, 0.040)** |  | **0.017** |
| **Model 1** | Regular (ref.) | – | 0.111 | – |  | Regular (ref.) | – | 0.105 | – |
|  | Moderately regular | 0.012 (–0.008, 0.032) |  | 0.380 |  | Moderately regular | –0.004 (–0.022, 0.013) |  | 0.625 |
|  | Highly irregular | 0.020 (0.000, 0.040) |  | 0.174 |  | Highly irregular | –0.004 (–0.022, 0.013) |  | 0.783 |
| **Model 2A** | Regular (ref.) | – | 0.106 | – |  | Regular (ref.) | – | 0.105 | – |
|  | Moderately regular | 0.008 (–0.012, 0.028) |  | 0.350 |  | Moderately regular | –0.005 (–0.023, 0.013) |  | 0.595 |
|  | Highly irregular | 0.012 (–0.008, 0.032) |  | 0.210 |  | Highly irregular | –0.003 (–0.022, 0.015) |  | 0.712 |
| **Model 2B** | Regular (ref.) | – | 0.111 | – |  | Regular (ref.) | – | 0.105 | – |
|  | Moderately regular | 0.008 (–0.012, 0.028) |  | 0.350 |  | Moderately regular | –0.004 (–0.022, 0.013) |  | 0.619 |
|  | Highly irregular | **0.035 (0.017, 0.053)** |  | **<0.001** |  | Highly irregular | –0.003 (–0.021, 0.016) |  | 0.775 |

All models used log-transformed depression symptoms as the dependent variable. Model 1 adjusted for gender, marital status, education, smoking, heavy alcohol use, psychotropic medication use, chronotype, and work schedule. Models 2A and 2B were further adjusted for physical activity (A) or sedentary time (B).

Greater variability in sleep timing was associated with higher anxiety symptoms in the unadjusted model. Participants in the highly irregular group had significantly higher anxiety scores compared to those in the regular group (B = 0.042, 95% CI [0.024, 0.060], p < 0.001). Regarding sleep duration regularity, the unadjusted model likewise showed that highly irregular duration was associated with higher anxiety symptoms (B = 0.033, 95% CI [0.015, 0.051], p < 0.001). After adjusting for sociodemographic, lifestyle, and sleep-related covariates (Model 1), the association between sleep timing and depressive symptoms remained significant (B = 0.023, 95% CI [0.005, 0.041], p = 0.011). Further adjustment for physical activity (Model 2A) and sedentary time (Model 2B) yielded similar consistent associations (Model 2A: B = 0.025, 95% CI [0.007, 0.043], p = 0.001; Model 2B: B = 0.025, 95% CI [0.007, 0.043], p = 0.001; Table 3).

**Table 3.** The Association Between Sleep Midpoint Regularity/Sleep Duration Regularity and Anxiety Symptom Scores Among 46-year-old Northern Finland Birth Cohort 1966 Participants (n = 3,556) Using Multivariable Regression Analyses

| Model | Sleep timing regularity |  |  |  |  | Sleep duration regularity |  |  |  |
| --- | --- | --- | --- | --- | --- | --- | --- | --- | --- |
|  | Category | B (95% CI) | Adjusted R<sup>2</sup> | p |  | Category | B (95% CI) | Adjusted R<sup>2</sup> | p |
| Unadjusted | Regular (ref.) | – | 0.011 | – |  | Regular (ref.) | – | 0.004 | – |
|  | Moderately regular | 0.013 (-0.007, 0.033) |  | 0.280 |  | Moderately regular | 0.008 (-0.010, 0.026) |  | 0.373 |
|  | Highly irregular | **0.042 (0.024, 0.060)** |  | **<0.001** |  | Highly irregular | **0.033 (0.015, 0.051)** |  | **<0.001** |
| Model 1 | Regular (ref.) | – | 0.123 | – |  | Regular (ref.) | – | 0.121 | – |
|  | Moderately regular | 0.008 (-0.012, 0.028) |  | 0.340 |  | Moderately regular | 0.003 (0.014, 0.020) |  | 0.730 |
|  | Highly irregular | **0.023 (0.005, 0.041)** |  | **0.011** |  | Highly irregular | 0.006 (-0.011, 0.024) |  | 0.476 |
| Model 2A | Regular (ref.) | – | 0.130 | – |  | Regular (ref.) | – | 0.125 | – |
|  | Moderately regular | 0.005 (-0.015, 0.025) |  | 0.545 |  | Moderately regular | 0.001 (-0.016, 0.018) |  | 0.896 |
|  | Highly irregular | **0.025 (0.007, 0.043)** |  | **0.001** |  | Highly irregular | 0.002 (-0.016, 0.020) |  | 0.803 |
| Model 2B | Regular (ref.) | – | 0.127 | – |  | Regular (ref.) | – | 0.124 | – |
|  | Moderately regular | 0.007 (-0.013, 0.027) |  | 0.434 |  | Moderately regular | 0.002 (-0.015, 0.020) |  | 0.780 |
|  | Highly irregular | **0.025 (0.007, 0.043)** |  | **0.001** |  | Highly irregular | 0.006 (-0.012, 0.023) |  | 0.524 |

All models used log-transformed anxiety symptoms as the dependent variable. Model 1 adjusted for gender, marital status, education, smoking, heavy alcohol use, psychotropic medication use, chronotype, and work schedule. Models 2A and 2B were further adjusted for physical activity (A) or sedentary time (B).

We examined whether accelerometer-measured total physical activity and sedentary time moderate the associations between sleep regularity (sleep timing regularity and sleep duration regularity) and symptoms of anxiety and depression using fully adjusted linear regression models with interaction terms (Tables 4 and 5). Neither physical activity nor sedentary time significantly moderated these associations (all interaction p > 0.2). Main effects showed that a highly irregular sleep timing was associated with higher anxiety scores (B = 0.021, 95% CI [0.003, 0.038], p = 0.023), and highly irregular sleep duration showed a similar association with higher anxiety scores (B = 0.020, 95% CI [0.002, 0.037], p = 0.030). Physical activity had an independent protective association with anxiety (p = 0.048), but not with depression (p = 0.584; Table 4).

**Table 4.**
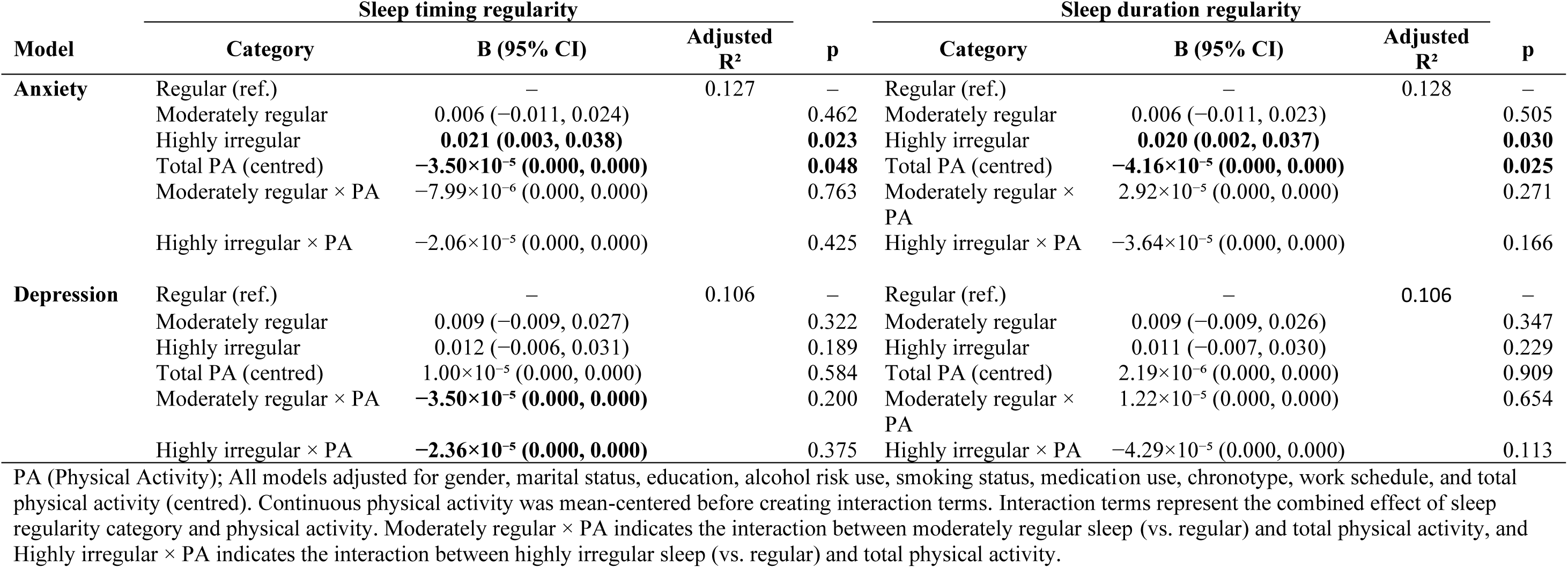
Moderation Analyses by Total Physical Activity on the Association Between Sleep Timing Regularity/Sleep Duration Regularity and HSCL-25 Anxiety and Depression Symptoms Among 46-year-old Northern Finland Birth Cohort 1966 Participants (n = 3,556) by Multivariable Regression Analyses

**Table 5.** Moderation Analyses by Sedentary Time on the Association Between Sleep Timing Regularity/Sleep Duration Regularity and HSCL-25 Anxiety and Depression Symptoms Among 46-Year-Old Northern Finland Birth Cohort 1966 Participants (n = 3,556) by Multivariable Regression Analyses

|  | Sleep timing regularity |  |  |  |  | Sleep duration regularity |  |  |
| --- | --- | --- | --- | --- | --- | --- | --- | --- |
| Model | Category | B (95% CI) | Adjusted R<sup>2</sup> | p | Category | B (95% CI) | Adjusted R<sup>2</sup> | p |
| Anxiety | Regular (ref.) | — | 0.127 | — | Regular (ref.) | — | 0.125 | — |
|  | Moderately regular | 0.007 (−0.010, 0.024) |  | 0.432 | Moderately regular | 0.007 (−0.010, 0.025) |  | 0.408 |
|  | Highly irregular | **0.020 (0.002, 0.038)** |  | **0.030** | Highly irregular | **0.021 (0.003, 0.039)** |  | **0.023** |
|  | Sedentary time (centred) | 0.000 (0.000, 0.000) |  | 0.052 | Sedentary time (centred) | 9.40×10<sup>−5</sup> (0.000, 0.000) |  | 0.188 |
|  | Moderately regular × Sedentary time | 0.000 (0.000, 0.000) |  | 0.231 | Moderately regular × Sedentary time | −3.22×10<sup>−5</sup> (0.000, 0.000) |  | 0.746 |
|  | Highly irregular× Sedentary time | 0.000 (0.000, 0.000) |  | 0.517 | Highly irregular× Sedentary time | 9.87×10<sup>−5</sup> (0.000, 0.000) |  | 0.313 |
| Depression | Regular (ref.) | — | 0.105 |  | Regular (ref.) | — | 0.106 |  |
|  | Moderately regular | 0.009 (−0.009, 0.027) |  | 0.310 | Moderately regular | 0.009 (−0.008, 0.027) |  | 0.298 |
|  | Highly irregular | 0.009 (−0.009, 0.027) |  | 0.199 | Highly irregular | 0.012 (−0.006, 0.031) |  | 0.187 |
|  | Sedentary time (centred) | −3.15×10<sup>−5</sup> (0.000, 0.000) |  | 0.651 | Sedentary time (centred) | −5.04×10<sup>−5</sup> (0.000, 0.000) |  | 0.493 |
|  | Moderately regular × Sedentary time | 5.1×10<sup>−5</sup> (0.000, 0.000) |  | 0.619 | Moderately regular × Sedentary time | 5.18×10<sup>−5</sup> (0.000, 0.000) |  | 0.613 |
|  | Highly irregular× Sedentary time | 8.47×10<sup>−5</sup> (0.000, 0.000) |  | 0.393 | Highly irregular× Sedentary time | 0.000 (0.000, 0.000) |  | 0.191 |

All models adjusted for gender, marital status, education, alcohol risk use, smoking status, medication use, chronotype, work schedule, and total sedentary time (centred). Continuous physical activity was mean-centered before creating interaction terms. Interaction terms represent the combined effect of sleep regularity category and physical activity. Moderately regular × Sedentary time indicates the interaction between moderately regular sleep (vs. regular) and total physical activity, and Highly irregular × Sedentary time indicates the interaction between highly irregular sleep (vs. regular) and total physical activity.

Regarding anxiety symptoms, a highly irregular midpoint of time in bed was significantly associated with increased anxiety scores (B = 0.020, 95% CI [0.002, 0.038], p = 0.030), while the moderately regular group showed no significant effect. Furthermore, highly irregular bed duration times remained significantly associated with higher anxiety symptoms even after controlling for sedentary time (B = 0.021, 95% CI [0.003, 0.039], p = 0.023). No significant interaction was found between sedentary time and time in bed regularity for either anxiety or depression symptoms. Sedentary time was not significantly associated with anxiety or depression (Table 5).

## Discussion

In this study of 3,556 middle-aged adults, sleep timing irregularity (variability in the midpoint of the sleep period) was consistently linked to higher anxiety symptoms. Associations with depressive symptoms disappeared when physical activity was considered as a factor, but the association was re-established when sedentary time was examined instead. These findings suggest that physical activity may partly explain the link between sleep timing irregularity and depressive symptoms, whereas associations with anxiety appear more independent of movement behaviors. Variability in sleep duration did not show stable associations with anxiety or depression symptoms. These results indicate that timing-based sleep metrics show clearer associations with midlife anxiety than variability in sleep duration.

These findings align with previous research demonstrating that greater sleep-timing irregularity is linked to poorer mental health, particularly anxiety. In the UK Biobank study of adults, sleep regularity was quantified using the Sleep Regularity Index (SRI) computed from 7-day wrist-accelerometry data; the study found that a higher SRI prospectively predicted lower incident anxiety and depression (Li et al., 2025). In adolescents, SRI computed from ∼6-month actigraphy was similarly related to fewer depressive symptoms (Castiglione-Fontanellaz et al., 2023). By contrast, in our midlife cohort, depression’s association with sleep-timing irregularity was not clear after adjustment, whereas anxiety’s association was more consistent. Notably, our exposure captured midpoint-of-sleep variability (timing), which differs from SRI’s minute-level regularity. Our study’s data were also captured over a shorter period than the adolescent monitoring window in the study by Castiglione-Fontanellaz et al. (2023).

This study’s findings align with prior research indicating that variability in accelerometer-determined sleep duration is generally less predictive of mental health outcomes than timing irregularity (Swanson et al., 2023). Age-related differences may partly explain these discrepancies, as older adults with depression have been shown to have irregular sleep–wake patterns (Luik et al., 2015), while middle-aged adults may be buffered by greater psychosocial stability. Observational research indicates that greater sleep regularity is associated with fewer depressive symptoms (Lunsford-Avery et al., 2018), although the exact role of sleep variability remains unclear. Broader evidence suggests that improving sleep and circadian health may benefit depression and anxiety (Freeman et al., 2020), but this does not specifically describe sleep regularity’s role.

We found no evidence that physical activity or sedentary time moderated the associations between sleep regularity and symptoms of anxiety or depression. Instead, irregular sleep timing was consistently associated with higher anxiety symptoms, independent of other lifestyle factors. Physical activity showed an independent protective association with anxiety, whereas sedentary time was not significantly related to anxiety or depression. These findings align with previous meta-analyses and longitudinal studies demonstrating the protective role of physical activity against anxiety and depression (Kandola et al., 2021; Schuch et al., 2018) and support recent evidence that replacing sedentary time with physical activity may mitigate adverse mental health effects (Zhu et al., 2024). Recent research has shown that irregular sleep timing combined with lower physical activity or increased sedentary time is associated with increased depressive symptoms (Kang et al., 2024; Lewis et al., 2021). Large-scale evidence also indicates that sleep and physical activity have independent and joint associations with mental health (Brown et al., 2025), supporting the relevance of integrating lifestyle factors when examining sleep patterns. Additionally, accelerometer-derived circadian rhythm disruption (indexed by lower rest–activity relative amplitude) has been associated with higher odds of mood disorders, including depression (Lyall et al., 2018).

This study benefits from several key methodological strengths that improve the reliability and validity of its findings. First, it is among the few studies to investigate the association between accelerometer-determined sleep regularity and mental distress symptoms in a large, population-based cohort of middle-aged adults, a demographic often underrepresented in sleep and mental health research. Sleep behavior was measured using Polar Active wrist-worn accelerometers over 7 consecutive days, capturing sleep periods, physical activity, and sedentary time, thereby reducing recall bias associated with self-reports (Cespedes et al., 2016; Lauderdale et al., 2008). The large sample (n = 3,556) from the well-characterized NFBC1966 cohort improved statistical power and generalizability. Furthermore, the comprehensive adjustment for a wide range of sociodemographic, lifestyle, work-related, and chronobiological covariates strengthens the internal validity of the findings. The prevalence of clinically relevant symptoms in our sample was relatively low compared to clinical populations, but it aligns with general population estimates. Specifically, 9.6% of participants reported anxiety symptoms and 11.8% reported depressive symptoms, which is consistent with population-based rates (Kessler et al., 2012) and substantially lower than those observed in clinical samples (Baxter et al., 2013; Tinghög et al., 2017). These findings suggest that our cohort reflects a generally healthy working-age population. While the exclusion of participants with missing HSCL-25 data and listwise deletion of covariates with < 5% missingness may have marginally affected representativeness, the large sample size and objective measures support reliability.

Several limitations should be acknowledged. The cross-sectional design precludes causal inference, and a bidirectional relationship between sleep and mental health has been previously well-documented in longitudinal studies (Alvaro et al., 2013; James et al., 2018; Jansson-Fröjmark & Lindblom, 2008). Residual confounding may persist from unstudied factors such as undiagnosed sleep disorders, chronic conditions, or stress. Although HSCL-25 is validated for symptom assessment, it does not provide clinical diagnoses and may be influenced by individual response styles. Generalizability is limited by the cohort’s homogeneity (predominantly female, age 46) and cultural context. Sleep regularity was derived from a 7-day consecutive monitoring period, which may not fully capture long-term habitual patterns, although prior evidence suggests that such a measurement period generally provides a reliable estimate of habitual sleep (Lau et al., 2022).

In this population-based study of middle-aged adults, irregular sleep timing, assessed by variability in the midpoint of the sleep period, showed a stronger association with anxiety symptoms than variability in sleep period duration. In addition, physical activity and sedentary time did not moderate the relationship between sleep regularity and symptoms of anxiety or depression. These findings underscore the relevance of sleep timing regularity as a potential target for anxiety management in midlife and indicate a need for longitudinal research to confirm these associations and clarify underlying mechanisms.

## Data Availability

NFBC data is available from the University of Oulu, Infrastructure for Population Studies. Permission to use the data can be applied for research purposes via electronic material request portal. In the use of data, we follow the EU general data protection regulation (679/2016) and Finnish Data Protection Act. The use of personal data is based on cohort participant's written informed consent at his/her latest follow-up study, which may cause limitations to its use. Please, contact NFBC project center (NFBCprojectcenter(at)oulu.fi) and visit the cohort website for more information.

## Declarations

### Ethics approval and consent to participate

The 46-year follow-up study was approved by the Ethical Committee of the Northern Ostrobothnia Hospital District in Oulu, Finland (94/2011). The participants and their parents provided written consent for the 1966 study.

### Consent for publication

The study is part of the Northern Finland Birth Cohort 2015. Consent for publication was obtained on 5 January 2026.

### CRediT authorship contribution statement

SC: Investigation; Data curation; Formal analysis; Writing – original draft; Writing – review & editing. LN: Conceptualization; Methodology; Investigation; Data curation; Formal analysis; Writing – original draft; Writing – review & editing. MN: Methodology; Investigation; Data curation; Funding acquisition; Writing – review & editing. RK: Investigation; Funding acquisition; Writing – review & editing. VF: Conceptualization; Methodology; Investigation; Funding acquisition; Writing – review & editing.

### Funding

The NFBC1966 received financial support from the University of Oulu Grant No. 24000692, the University of Oulu Hospital Grant No. 24301140, and the ERDF European Regional Development Fund Grant No. 539/2010 A31592. The study was financially supported by the Ministry of Education and Culture, Finland (grant numbers OKM/86/626/2014, OKM/43/626/2015, OKM/17/626/2016, OKM/54/626/2019, OKM/85/626/2019, OKM/1096/626/2020, OKM/20/626/2022, and OKM/76/626/2024). MN has received funding from The University of Oulu & The Research Council of Finland (336449, 361560).

## Acknowledgments

We thank all cohort members and researchers who participated in the 46-year follow-up study. We also wish to acknowledge the work of the NFBC project center.

## Competing interests

Financial Disclosure: None. Non-Financial Disclosure: None.

## Notes

### Competing Interest Statement

The authors have declared no competing interest.

